# Validation of Estimated Glomerular Filtration Slope and Proteinuria Reduction as Surrogate Endpoints in IgA Nephropathy: A post-hoc Analysis from the J-IGACS

**DOI:** 10.1101/2025.06.15.25329660

**Authors:** Takaya Sasaki, Nobuo Tsuboi, Kentaro Koike, Hiroyuki Ueda, Masahiro Okabe, Shinya Yokote, Akihiro Shimizu, Keita Hirano, Tetsuya Kawamura, Takashi Yokoo, Yusuke Suzuki, the J-IGACS working group

## Abstract

**Background:** Estimated glomerular filtration rate (eGFR) slope and proteinuria reduction have been proposed as surrogate endpoints for kidney outcomes in IgA nephropathy (IgAN), but their validity remains under debate. We aimed to evaluate the surrogate potential of these markers in the context of corticosteroid therapy within a large Japanese cohort.

**Methods:** Patients with biopsy-proven IgAN from the Japan IgA Nephropathy Cohort Study (J-IGACS) were analyzed. Patients were categorized based on corticosteroid exposure within 12 months of diagnosis. To minimize confounding, overlap weighting was used to balance baseline characteristics. The primary outcome was a composite kidney endpoint defined as a ≥40 decline in eGFR or the initiation of kidney replacement therapy. Secondary outcomes included 2-year eGFR slope and proteinuria ratio at 1-year.

**Results:** Corticosteroid therapy was associated with a lower incidence of the composite kidney outcome (15.5 vs. 28.2%, P = 0.007), slower decline in eGFR (−0.30 vs. −2.99 mL/min/1.73 m^2^/year, P = 0.001), and lower proteinuria ratio (0.521 vs. 0.235, P < 0.001). These associations were consistent with predictions from established meta-regression models that linked changes in surrogate markers to kidney outcomes across diverse kidney diseases including IgAN.

**Conclusion:** Both the eGFR slope and proteinuria ratio demonstrated strong and consistent associations with kidney outcomes in the context of corticosteroid therapy. These findings support their validity as surrogate endpoints for clinical trials and suggest their usefulness for risk stratification and therapeutic monitoring in routine nephrology practice.

**Lay-Summary:** IgA nephropathy (IgAN) is a common kidney disease that can lead to kidney failure over time. To monitor treatment effectiveness, researchers often use early indicators like how quickly kidney function declines (eGFR slope) and changes in protein levels in urine (proteinuria). However, whether these markers truly reflect long-term outcomes remains uncertain. In this study, we analyzed over 1,000 patients with biopsy-confirmed IgAN from a nationwide Japanese cohort. We compared those who received corticosteroid treatment within a year of diagnosis to those who did not. After adjusting for differences between groups, we found that corticosteroid use was linked to better kidney outcomes, slower decline in kidney function, and reduced proteinuria. These results matched predictions from large-scale international studies. Our findings support the use of eGFR slope and proteinuria as reliable surrogate endpoints in clinical trials and everyday care, helping doctors assess treatment response more quickly and accurately.

## Introduction

IgA nephropathy (IgAN) is the most prevalent form of primary glomerulonephritis globally and a major cause of chronic kidney disease (CKD) and kidney failure [1, 2]. Because IgAN progresses slowly over many years [3–5], establishing reliable surrogate endpoints that can be evaluated over a short period is crucial for improving the efficiency of clinical trials and guiding personalized treatment strategies as new therapies emerge. Among candidate surrogate markers, the trajectory of the estimated glomerular filtration rate (eGFR) slope and reduction in proteinuria have been proposed [6–9]. However, their validity remains a topic of active discussion, particularly when applied to therapeutic interventions.

Corticosteroid therapy has long been a cornerstone of treatment for IgAN, particularly in patients with persistent proteinuria despite supportive care. Importantly, randomized controlled trials [10] and meta-analysis [11] studies have consistently demonstrated that corticosteroids significantly reduce proteinuria and slow disease progression, establishing their efficacy as a key therapeutic option. In Japan, in particular, corticosteroid therapy has become widely adopted as a standard treatment in routine clinical practice [12].

Previous meta-regression analyses across various CKDs, including IgAN, have suggested that treatment-induced improvements in eGFR slope and reductions in proteinuria are associated with favorable kidney outcomes [9, 13–15]. Nevertheless, extrapolation of these findings to IgAN, especially in real-world cohorts with heterogeneous treatment responses, requires careful evaluation. To address these gaps, we leveraged data from the Japan IgA Nephropathy Cohort Study (J-IGACS) [12, 16] and focused on corticosteroid therapy as a representative treatment intervention. Using overlap weighting to emulate randomization and balance baseline characteristics, we examined whether corticosteroid exposure was associated with improvements in eGFR slope, reduction in proteinuria, and attenuation of kidney disease progression. In addition, we evaluated the consistency between observed treatment effects and predictions derived from established meta-regression models [9, 14, 15]. Through this approach, we aimed to validate eGFR slope and proteinuria reduction as robust surrogate endpoints for kidney outcomes in IgAN, with particular emphasis on their applicability in clinical trials and real-world practice.

## Methods

### Study Design and Population

This study is a post hoc analysis derived from the Japan IgA Nephropathy Cohort Study (J-IGACS), a prospective, multicenter registry enrolling individuals with biopsy-proven primary IgAN from April 1, 2005, to August 31, 2015 [12, 16]. Among the inclusion criteria for the original J-IGACS study was a diagnostic kidney biopsy containing a minimum of 10 glomeruli, a threshold chosen to ensure adequate histological assessment. For this analysis, we included 706 of the 991 patients from the primary study, excluding those with missing covariates. The present study was designed to confirm that treatment effects (as measured by the hazard ratio [HR], annual eGFR slope, and proteinuria ratio) derived from propensity score-based weighting were consistent across outcomes, as shown in **Figure 1**. Moreover, we checked the consistency between the HR observed in this study and HRs estimated from existing meta-regression equations. Ethical approval was granted by The Jikei University School of Medicine (approval no. 37-069 [12706]). All procedures conformed to the ethical principles outlined in the Declaration of Helsinki and adhered to the STROBE reporting standards.

**Figure 1.**
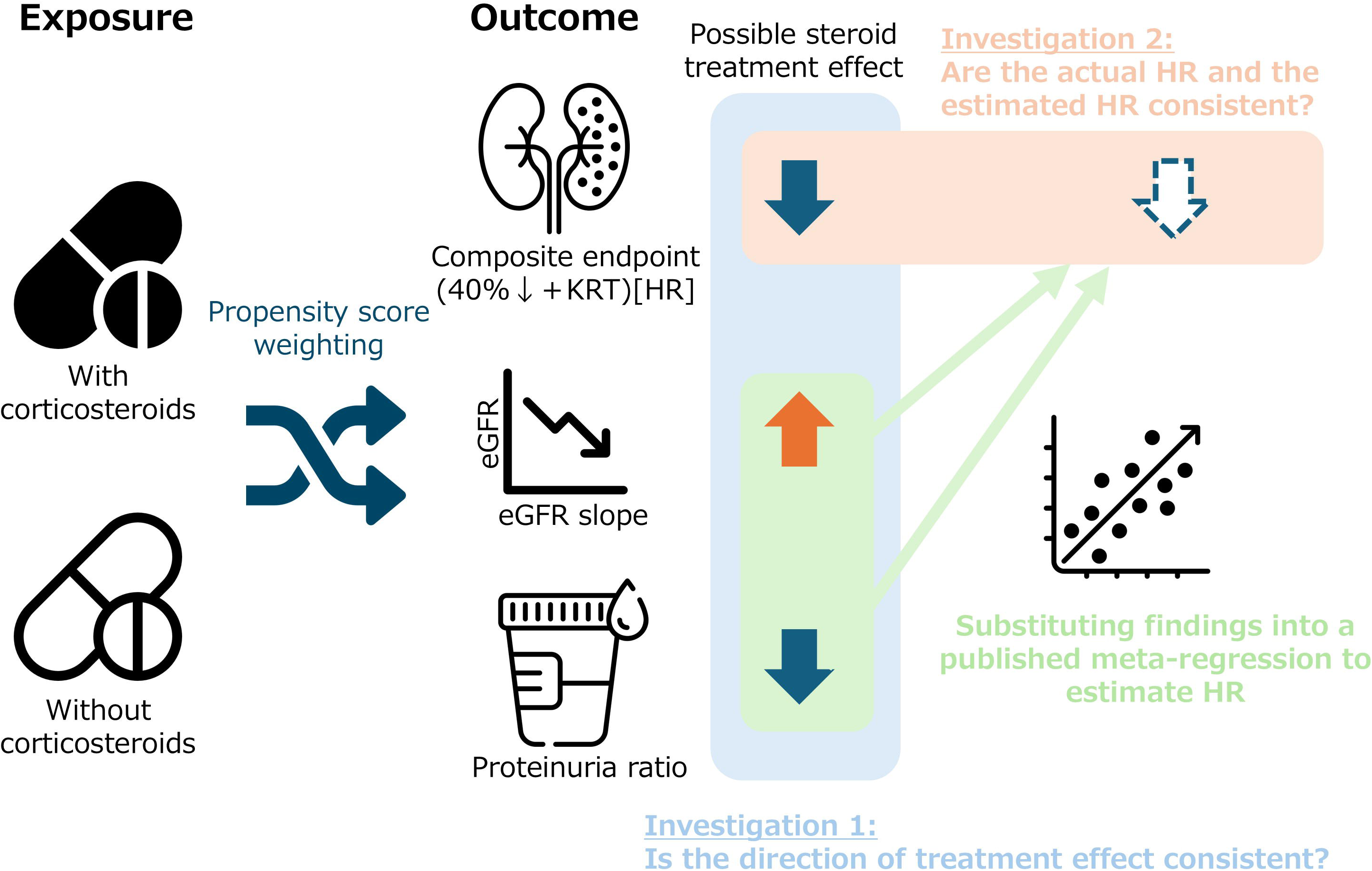
Overview of the study design and analytical approach. The following outcomes were calculated and compared between corticosteroid-treated and -untreated groups, with overlap weighting used to balance baseline characteristics: the hazard ratio (HR) for the composite kidney endpoint, defined as a ≥40% decline in estimated glomerular filtration rate (eGFR) or initiation of kidney replacement therapy (KRT); the annual eGFR slope over a 2-year period; and the proteinuria ratio at 1 year relative to baseline. In addition, the values observed in this study were compared with estimates based on meta-regression equations.

### Data Acquisition and Definitions

Demographic and clinical parameters, including age, sex, mean arterial pressure (MAP), serum creatinine, estimated glomerular filtration rate (eGFR), serum uric acid, 24-hour urinary proteinuria excretion (the urinary protein-to-creatinine ratio [UPCR] was permitted in place of 24-hour urinary protein excretion at all time points except baseline), and urinary red blood cell count, were systematically recorded at baseline and at 6-month intervals thereafter. eGFR was determined using Uemura’s equation [17] for participants under 20 years and Matsuo’s formula [18] for those aged 20 years and older. Histopathological assessment was based on the Oxford MEST-C classification criteria [19, 20]. Therapeutic interventions administered within 12 months of biopsy, including corticosteroids,renin-angiotensin-aldosterone system (RAAS) inhibitors, and tonsillectomy, were documented.

### Exposure Definition and Propensity Score-based Weighting Approach

Participants were stratified according to corticosteroid therapy initiation within one year following biopsy. To address baseline imbalances between treatment and non-treatment groups, propensity scores were calculated using logistic analysis including treatment as a dependent variable based on baseline covariates as independent variables, and overlap weighting was applied. Each participant was weighted by the probability of belonging to the opposite treatment group, emphasizing individuals with substantial covariate overlap.

### Endpoints

For kidney outcomes, the primary endpoint was a composite of 40% or greater decline in eGFR from baseline based on two consecutive measurements or initiation of kidney replacement therapy. Secondary outcomes included (i) the 2-year eGFR slope, calculated from all available eGFR values from baseline to 2 years using a linear mixed model for repeated measures, and (ii) proteinuria reduction assessed using the proteinuria ratio at 1 year, defined as 1-year over baseline proteinuria.

### Hazard Ratio and eGFR Slope Estimated from Meta-regression

To estimate the expected treatment effects on kidney outcomes, previously published meta-regression models were applied to the observed changes in surrogate markers.

The observed treatment effect on annual eGFR slope was substituted into two different meta-regression equations that relate eGFR slope to the hazard ratio (HR) for kidney outcomes. The first equation was derived from randomized controlled trials in IgAN, where eGFR slope was calculated over a 1-year period [9]. The second equation was developed from trials in broader CKD populations, using slopes calculated over at least 2 years [15].

The latter was included to reflect current methodological recommendations for slope-based analyses, which favor longer follow-up to ensure stable estimates.

The effect of corticosteroid treatment on the proteinuria ratio was entered into a separate meta-regression equation—derived from IgA nephropathy trials—that links the degree of proteinuria reduction to the HR for kidney outcomes [9]. This model was used to estimate the expected HR based solely on the proteinuria-lowering effect of treatment.

In addition, the observed proteinuria ratio was substituted into a meta-regression equation to estimate eGFR slope [14]. This allowed indirect estimation of slope effects when proteinuria was used as the primary marker of treatment response.

All estimated values from these models were then visually compared to the observed values in the current study using forest plots.

### Statistical Analyses

Descriptive statistics were applied to summarize baseline characteristics, expressed as mean (standard deviation), median [interquartile range], or frequencies and percentages, as appropriate. Weighted standardized differences were calculated to assess balance between groups. Cox proportional hazards models were fitted using overlap weights based on propensity scores, and baseline survival functions were subsequently estimated to generate weighted survival curves based on corticosteroid treatments. Cox proportional hazards models incorporating overlap weights were also used to estimate HRs with 95% confidence intervals. Differences in annual eGFR slopes according to corticosteroid exposure were evaluated using a linear mixed-effects model for repeated measures with random intercepts and slopes, incorporating all available eGFR measurements from the first two years.

Differences in the log-transformed proteinuria ratio between corticosteroid groups were assessed using a linear regression model. A two-sided P-value less than 0.05 was considered statistically significant. Statistical analyses were performed using SAS version 9.4 (SAS Institute Inc., Cary, NC, USA).

## Results

### Baseline Characteristics

Of the 991 patients in the primary study, a total of 706 patients with biopsy-confirmed IgAN and no missing covariates were included in the present analysis. Among them, 359 patients (50.8%) received corticosteroid therapy within 12 months of diagnosis. Before weighting, patients receiving corticosteroids had higher proteinuria levels and lower baseline eGFR compared to those not receiving corticosteroids. After applying overlap weighting, the baseline characteristics were well-balanced between the two groups, with weighted standardized differences of less than 0.001 for all covariates (**Table 1**).

**Table 1.** Baseline characteristics based on corticosteroid treatment before and after overlap weighting.

|  | Before weighting |  |  | After weighting |  |  |
| --- | --- | --- | --- | --- | --- | --- |
|  | Corticosteroid treatment |  | \|SMD\| | Corticosteroid treatment |  | \|SMD\| |
|  | Without<br>(n=230) | With<br>(n=476) |  | Without<br>(n=93.6) | With<br>(n=93.6) |  |
| **Basic information** |  |  |  |  |  |  |
| Age, year | 44.4 (17.1) | 37.1 (15.5) | 0.45 | 41.6 (19.2) | 41.6 (23.4) | <0.001 |
| Male sex, % | 49.6 | 52.1 | 0.05 | 50.6 | 50.6 | <0.001 |
| Weight, kg | 59.4 (12.5) | 58.8 (13.4) | 0.05 | 59.2 (16.6) | 59.2 (16.5) | <0.001 |
| Height, cm | 162.1 (9.8) | 162.1 (11) | 0.001 | 162.4<br>(10.9) | 162.4<br>(11.8) | <0.001 |
| **Past medical history** |  |  |  |  |  |  |
| Diabetes mellitus, % | 0.9 | 2.1 | 0.10 | 1.5 | 1.5 | <0.001 |
| Systolic blood pressure, mmHg | 125.1 (18) | 120.7<br>(16.9) | 0.25 | 123.4<br>(21.5) | 123.4<br>(25.3) | <0.001 |
| Antihypertensive drug, % | 43.5 | 37.4 | 0.12 | 39.0 | 39.0 | <0.001 |
| Statin, % | 12.2 | 9.9 | 0.07 | 11.0 | 11.0 | <0.001 |
| **Kidney-related measurements** |  |  |  |  |  |  |
| eGFR, mL/min/1.73 m<sup>2</sup> | 72.3 (27.9) | 77.1 (28.4) | 0.17 | 74.1 (32.9) | 74.1 (36.6) | <0.001 |
| UPE, g/day or g/g | 0.8 (1.4) | 1.4 (2.8) | 0.26 | 0.9 (2.5) | 0.9 (1.2) | <0.001 |
| Microhematuria, % | 99.6 | 99.6 | 0.002 | 99.6 | 99.6 | <0.001 |
| **Histopathological findings** |  |  |  |  |  |  |
| M1, % | 22.6 | 31.3 | 0.2 | 25.5 | 25.5 | <0.001 |
| E1, % | 21.7 | 42.6 | 0.46 | 30.7 | 30.7 | <0.001 |
| S1, % | 63.5 | 80.7 | 0.39 | 72.7 | 72.7 | <0.001 |
| T1 or 2, % | 23.9 | 21.2 | 0.06 | 21.7 | 21.7 | <0.001 |
| C1 or 2, % | 20.4 | 48.1 | 0.61 | 31.1 | 31.1 | <0.001 |
| **Blood tests** |  |  |  |  |  |  |
| Sodium, mmol/L | 140.4 (2.3) | 140.3 (2.1) | 0.09 | 140.2 (2.3) | 140.2 (3.0) | <0.001 |
| Potassium, mmol/L | 4.2 (0.4) | 4.2 (0.4) | 0.02 | 4.2 (0.4) | 4.2 (0.5) | <0.001 |
| ALT, IU/L | 19.0 (14.4) | 17.7 (13.1) | 0.10 | 18.0 (13.1) | 18.0 (15.3) | <0.001 |
| AST, IU/L | 20.6 (12.3) | 20.4 (10.2) | 0.01 | 20.5 (20.7) | 20.5 (12.0) | <0.001 |
| Alb, g/dL | 4.0 (0.4) | 3.9 (0.6) | 0.36 | 4.0 (0.6) | 4.0 (0.6) | <0.001 |
| Uric acid, mg/dL | 5.9 (1.7) | 5.8 (1.6) | 0.09 | 5.9 (1.8) | 5.9 (2.4) | <0.001 |
| IgA, g/L | 3.25 (1.20) | 3.42 (1.27) | 0.14 | 3.34 (1.68) | 3.34 (1.77) | <0.001 |
| Hb, g/dL | 13.4 (1.7) | 13.3 (1.7) | 0.05 | 13.3 (1.9) | 13.3 (2.4) | <0.001 |
| WBC, /μL | 6623<br>(5671) | 6656<br>(1838) | 0.01 | 6580<br>(2980) | 6580<br>(2650) | <0.001 |
| **Initial treatment** |  |  |  |  |  |  |
| Tonsillectomy, % | 13.0 | 59.2 | 1.10 | 25.9 | 25.9 | <0.001 |
| Immunosuppressive agent, % | 1.3 | 7.4 | 0.30 | 1.9 | 1.9 | <0.001 |
| RAS inhibitor, % | 68.3 | 56.5 | 0.24 | 62.0 | 62.0 | <0.001 |

### Kidney Outcomes

During a median follow-up of 6.5 years, 147 patients (20.8%) developed the composite kidney endpoint. The risk of the primary endpoint was significantly lower in the corticosteroid group compared to the non-corticosteroid group (15.5% vs. 28.2%).

Overlap-weighted Cox proportional hazards analysis demonstrated that corticosteroid exposure was associated with a significantly reduced risk of the composite endpoint (weighted HR: 0.405; 95% CI: 0.211–0.777; P = 0.007) (**Figure 2**).

**Figure 2.**
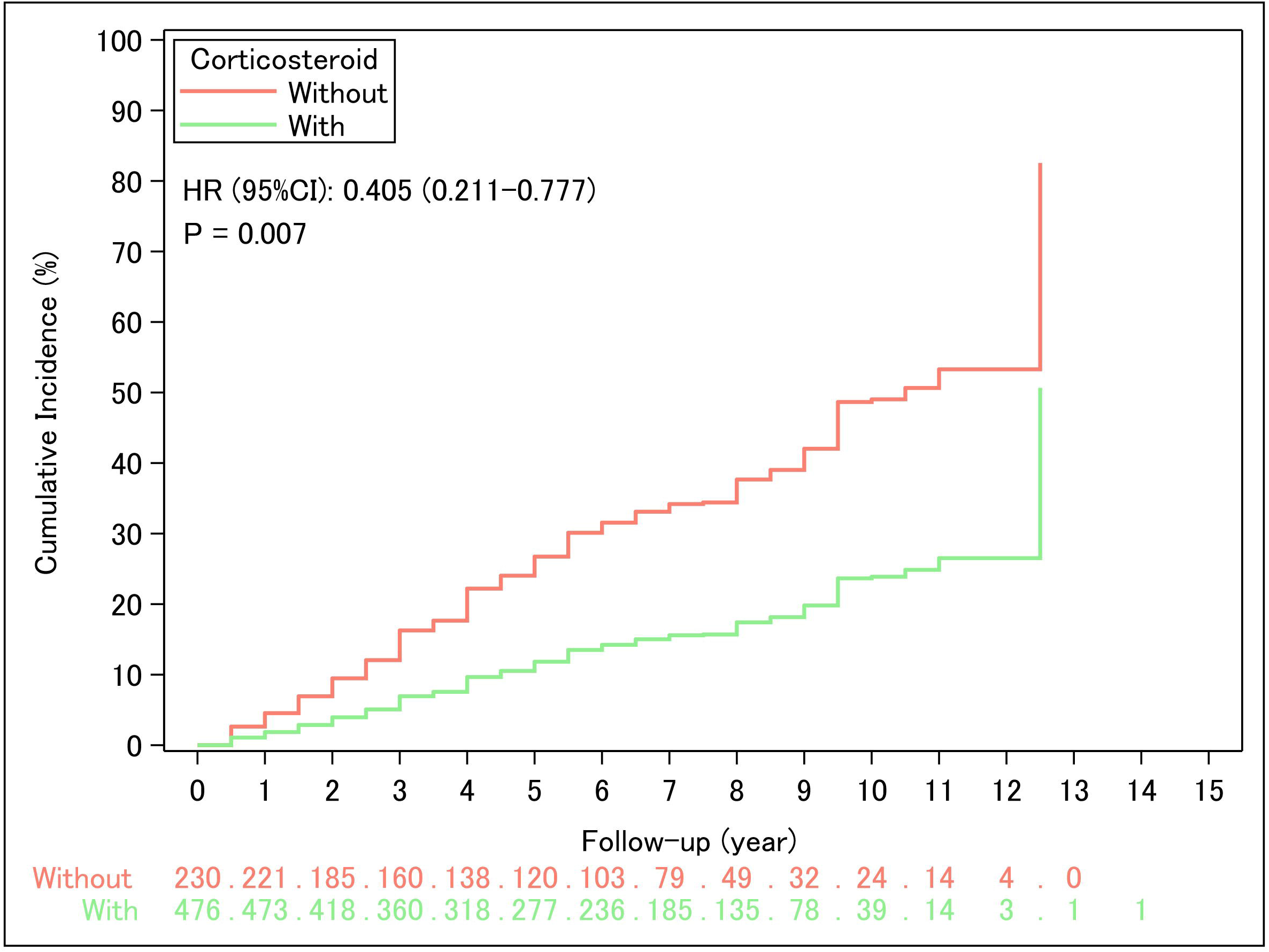
Weighted survival curves for the cumulative incidence of the composite kidney endpoint based on corticosteroid exposure. The composite endpoint was defined as ≥40% decline in estimated glomerular filtration rate or initiation of kidney replacement therapy. Cox proportional hazards models were fitted using overlap weighting, and baseline survival functions were subsequently estimated to generate weighted survival curves, and also used to estimate the hazard ratio (HR) with 95% confidence interval (CI).

### eGFR Slope

The mean eGFR decline over two years was significantly attenuated in patients receiving corticosteroids compared to those not receiving them (−0.30 vs. −2.99 mL/min/1.73 m^2^/year), corresponding to a between-group difference of +2.69 mL/min/1.73 m^2^/year (95%CI: +1.17–+4.21) (P = 0.01; **Figure 3**).

**Figure 3.**
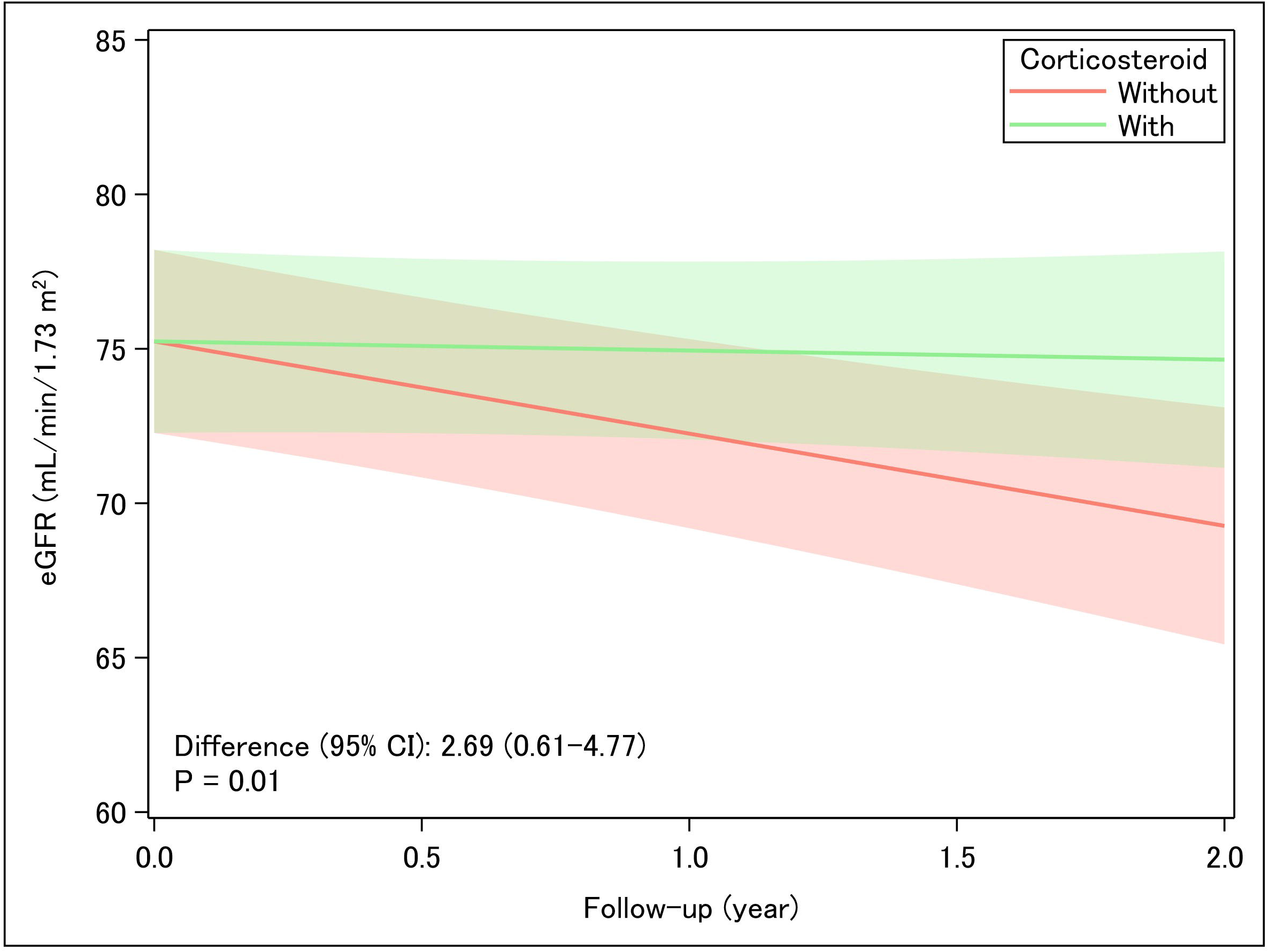
Comparison of the annual estimated glomerular filtration rate (eGFR) slope for 2 years between corticosteroid-treated and untreated groups after overlap weighting. A linear mixed-effect model for repeated measures with weighting was incorporated to depict the trajectory of eGFR over time.

### Proteinuria Ratio

Patients treated with corticosteroids showed a significantly greater reduction in proteinuria ratio at 1 year compared to those not receiving corticosteroids (0.521 vs 0.235) with a geometric mean difference of 0.452 (95%CI: 0.344–0.594) (P <0.001; **Figure 4**).

**Figure 4.**
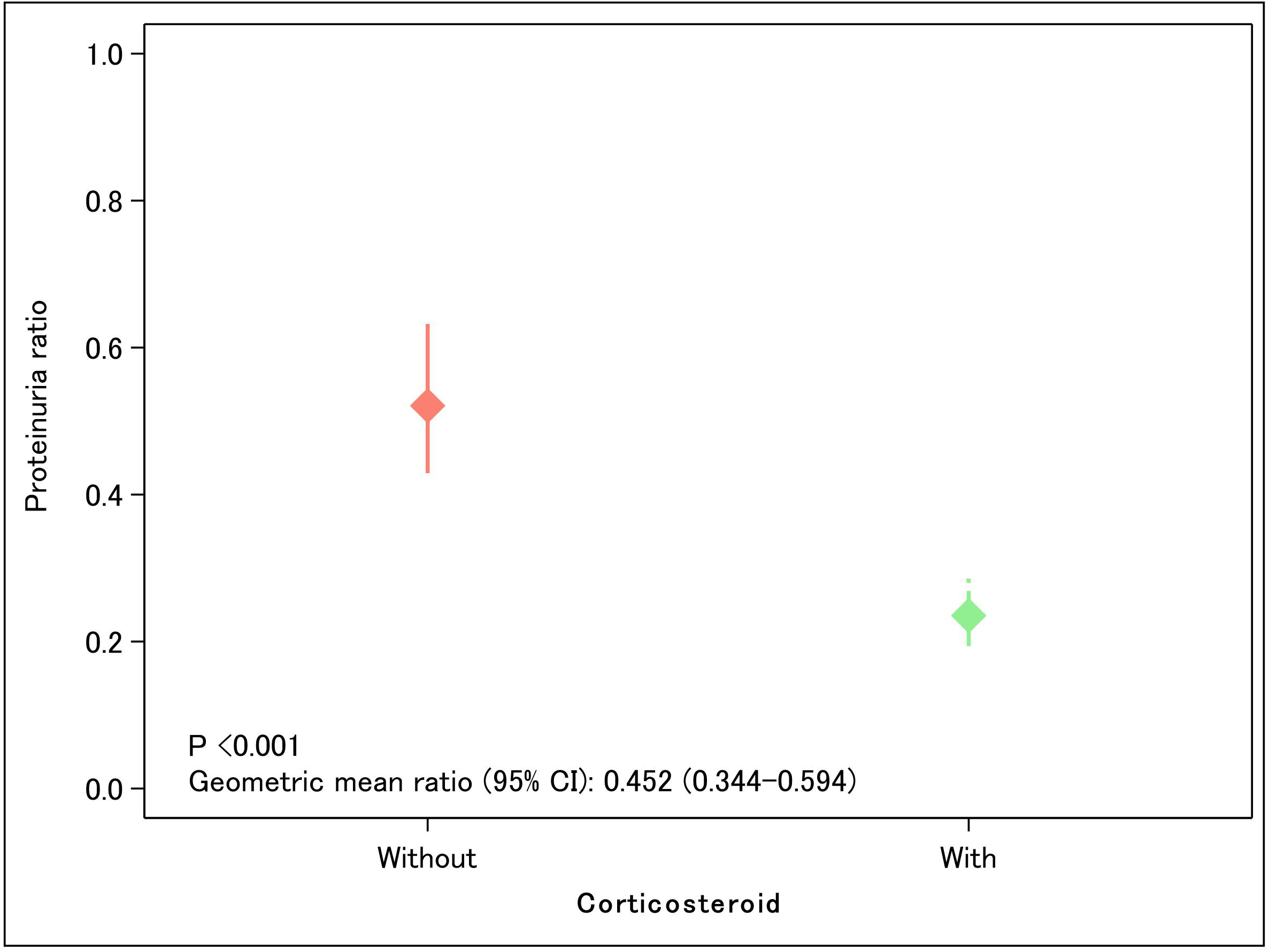
Comparison of proteinuria ratio at 1-year between corticosteroid-treated and -untreated groups after overlap weighting. A linear regression model with weighting was used for comparison.

### Comparison with Meta-Regression Estimations

To evaluate the validity of eGFR slope and proteinuria ratio as surrogate endpoints, we compared the actual observed effects of corticosteroid therapy on long-term kidney outcomes with the estimations derived from established meta-regression models. These models quantitatively link changes in surrogate markers to the risk of kidney disease progression based on aggregated data from multiple clinical trials. Specifically, the degree of proteinuria ratio and the improvement in the eGFR slope observed among corticosteroid-treated patients aligned closely with the magnitude of clinical benefit estimated by these models (**Figure 5**). The observed hazard reduction for the composite kidney outcome fell within the confidence intervals projected based on the changes in surrogate endpoints.

**Figure 5.**
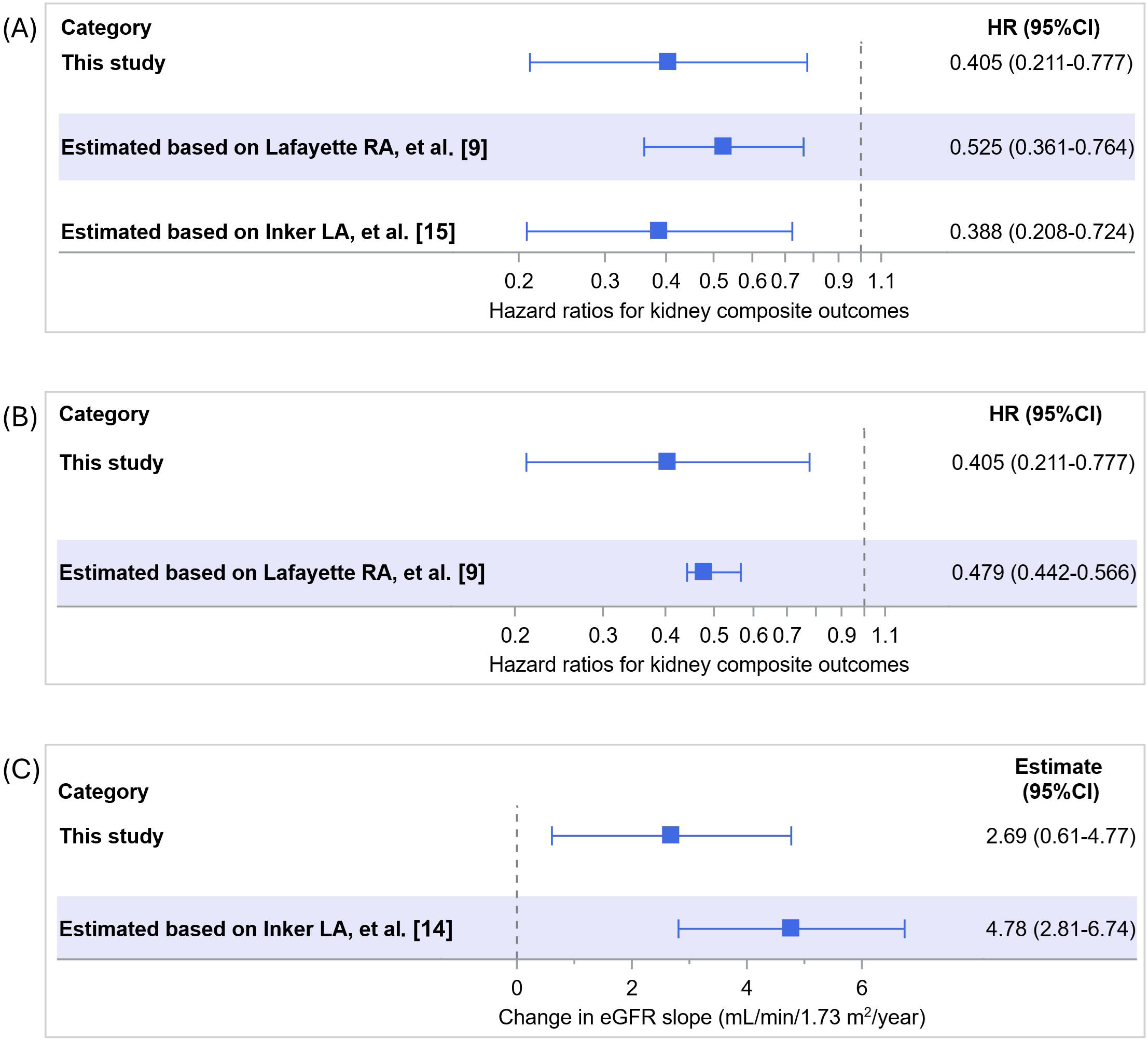
Observed versus estimated treatment effects based on meta-regression models linking surrogate endpoints to long-term kidney outcomes. (A) Observed hazard ratio (HR) and HR calculated from the estimated glomerular filtration rate (eGFR) slope. (B) Actual HR and HR calculated from proteinuria ratio. (C) Actual eGFR slope and eGFR estimated from proteinuria ratio.

## Discussion

In this post hoc analysis of a large-scale, prospective cohort of patients with biopsy-confirmed IgAN, we specifically focused on the evaluation of corticosteroid therapy as a key therapeutic intervention. By categorizing patients based on corticosteroid exposure and applying overlap weighting to achieve balance in baseline characteristics, we effectively minimized confounding by indication and approximated the conditions of a randomized controlled trial within a real-world setting. Our primary objective was to assess whether improvements in eGFR slope and proteinuria ratio, as surrogate markers, reliably reflected the clinical benefits of corticosteroid therapy. Importantly, the magnitude of the effects of treatment on both surrogate endpoints corresponded closely with the estimations derived from established meta-regression models. This consistency supports the validity of these markers as surrogate endpoints in the context of corticosteroid-induced therapeutic effects and underscores their utility for monitoring treatment response in clinical trials for IgAN.

Corticosteroid therapy is globally recognized for its efficacy in clinical trials and has long served as a cornerstone in the management of IgAN [10, 21]. In Japan, it has become firmly established as a standard treatment [12]. By focusing on this well-established intervention, our study provides critical clinically relevant insights into the utility of eGFR slope and proteinuria reduction as surrogate endpoints. Moreover, this approach enhances the translational impact of our study, as it reflects real-world treatment paradigms currently used in routine nephrology practice [22]. Our findings thus provide valuable insights into how surrogate markers perform not only in theoretical models but also in the context of established therapeutic strategies for IgAN.

Collectively, the present results reinforce the idea that early surrogate markers have clinical relevance in IgAN. Consistent with prior studies across various CKDs, including IgAN [9, 15], we found there that attenuation of eGFR decline and reductions in proteinuria [13, 14, 23] were associated with improved long-term kidney prognosis in patients with IgAN. Notably, the meta-regression models used for comparison were derived from aggregated data encompassing a wide range of therapeutic interventions, not limited to any specific treatment class. The consistency between the observed treatment effects of corticosteroid therapy and the model-based expectations thus suggests that improvements in these surrogate markers are reliably associated with clinical outcomes irrespective of the specific therapeutic strategy.

This finding further supports the validity of eGFR slope and proteinuria ratio as surrogate endpoints in IgAN, indicating that monitoring these parameters can meaningfully capture long-term therapeutic benefits across diverse treatment settings, including routine clinical practice. Together, these methodological strengths contribute to the rigor of our study and enhance confidence in the applicability of our findings to both clinical trial design and real-world clinical practice.

Some limitations should be acknowledged. This was a post hoc, observational analysis and residual confounding cannot be fully excluded despite rigorous adjustment. Although corticosteroid therapy was standardized to some extent, variations in dosing regimens, duration, and concurrent supportive treatments across institutions may have influenced treatment responses. In addition, the use of real-world clinical data, while enhancing generalizability, may have introduced variability in treatment protocols and outcome measurements. Finally, the generalizability of our findings to other ethnic groups remains to be confirmed, as the cohort was predominantly Japanese.

In conclusion, our study provides compelling evidence supporting the validity of eGFR slope and proteinuria ratio as surrogate endpoints for kidney outcomes in patients with IgAN receiving corticosteroid therapy. These markers may serve not only to facilitate endpoint evaluation in clinical trials but also to enhance risk stratification and therapeutic monitoring in routine nephrology practice.

## Authors’ Contributions

Conceptualization: Takaya Sasaki, Nobuo Tsuboi, Takashi Yokoo, Yusuke Suzuki.

Data curation: Takaya Sasaki.

Formal analysis: Takaya Sasaki.

Funding acquisition: Yusuke Suzuki.

Investigation: Takaya Sasaki, Nobuo Tsuboi.

Methodology: Takaya Sasaki.

Project administration: Nobuo Tsuboi, Takashi Yokoo, Yusuke Suzuki.

Supervision: Takashi Yokoo, Yusuke Suzuki.

Visualization: Takaya Sasaki.

Writing – original draft: Takaya Sasaki, Nobuo Tsuboi.

Writing – review & editing: Kentaro Koike, Hiroyuki Ueda, Masahiro Okabe, Shinya Yokote, Akihiro Shimizu, Keita Hirano, Tetsuya Kawamura, Takashi Yokoo, Yusuke Suzuki.

## Acknowledgments

This study was approved by the ethics review board of the Jikei University School of Medicine (approval no. 37-069 [12706]).

## Data availability statement

All data produced in the present study are available upon reasonable request to the authors.

